# Protocol for: Lockable Smartphone Pouches in UK Secondary Schools – A Cohort Study

**DOI:** 10.64898/2026.05.15.26353291

**Authors:** Jessica John, Anjali Khambhayta, Maximin Lange, Francesca Maher, Chloé Locatelli, Nicola Kalk, Ben Carter

**Author notes:** Corresponding author, Dr Maximin Lange, Address IoPPN, 16 De Crispingy Park, SE5, London. Joint First Author (Jessica John, Anjali Khambhayta). Joint Last Author (Nicola Kalk, Ben Carter).

## Abstract

**Introduction:** Smartphone ownership among UK adolescents is near universal, and teachers report phones increasingly being involved in classroom disruption, with misuse during school hours among the more common serious behavioural issues in secondary schools. Evidence on whether restrictive policies improve behaviour, attainment, or wellbeing remains limited. The primary objective is to assess the impact of a lockable smartphone pouch on educational attainment and behaviour. Secondary objectives are to assess impacts on general functioning, psychological wellbeing, and school-level indicators such as exclusions, and to examine whether effects differ for pupils who may be most at risk.

**Methods and analysis:** We will conduct a mixed methods cohort study in secondary schools across Northern Ireland and England during the 2025 to 2026 academic year. The quantitative component uses a serial cross-sectional design, with students completing an online questionnaire at 0, 4, and 8 weeks covering homework completion, classroom disruption, participation in PE and extracurricular activities, peer interaction, and smartphone use. Measures include the Strengths and Difficulties Questionnaire, the Revised Child Anxiety and Depression Scale, the short form of the Smartphone Addiction Scale, and the Bergen Social Media Addiction Scale. Schools will also supply half-termly aggregate data on exclusions, detentions, CAMHS referrals, counsellor visits, and parent visits from September 2023 to May 2026. Assuming 90% power, a two-sided type 1 error of 0.05, an intracluster correlation of 0.02, and 25% loss to follow up, we aim to recruit a minimum of 3,200 students from six or more schools to detect a small effect (Cohen’s d = 0.2) on SDQ hyperactivity score. Continuous outcomes will be analysed with linear regression and binary outcomes with logistic regression. Aggregate school data will be analysed using an interrupted time series design. Prespecified subgroup analyses cover SEN or neurodivergent status, area-level deprivation, and existing school phone policy. Qualitative data from focus groups with students and staff and semi-structured interviews with school leads will be analysed thematically using Braun and Clarke’s six-phase approach.

**Ethics and dissemination:** The study has been approved by the King’s College London Research Ethics Committee. A Data Protection Impact Assessment has been agreed with the Northern Ireland Department of Education. Findings will be disseminated through a final report to the Department of Education, peer-reviewed publications, conference presentations, and accessible summaries for participating schools, pupils, parents, and policy makers.

**Strengths and limitations of this study:**

- Mixed methods design combines individual-level survey data, school-level disciplinary records, and qualitative focus groups and interviews, allowing triangulation of pupil, staff, and institutional perspectives on a lockable smartphone pouch intervention.
- Use of validated instruments (SDQ, RCADS, SAS-SV, BSMAS) supports comparability with the wider literature on adolescent mental health and problematic smartphone use, and prespecified subgroup analyses allow examination of differential effects in pupils who may be most at risk.
- Inclusion of retrospective aggregate school data from September 2023 enables an interrupted time series analysis that strengthens causal inference beyond what a pre-post survey alone could provide.
- The serial cross-sectional design with follow-up at 0, 4, and 8 weeks limits the ability to detect longer-term effects on attainment, behaviour, and wellbeing.

## INTRODUCTION

Virtually all adolescents in the UK (99% of 12-to 15-year-olds) have internet access and spend time online daily, engaging in streaming, social media use, and messaging^1,2^.

Research on the association between children and young people’s health and time spent on a phone or online have gained in popularity^3–6^, but often come to conflicting and/or varying conclusions:

There is some evidence for a beneficial relationship between phone time and wellbeing, mostly at shorter durations^7–10^, and some find that phones can act as educational tools^11–14^.

At the same time, there is a growing concern regarding the impact of smartphone use on behaviour, health, and classroom management of pupils^15–17^. Mobile phones and digital technologies are implicated in disruptive incidents and negative learning outcomes during school hours^18–20^. Teachers describe inappropriate phone use during lessons, misuse of digital platforms, and incidents involving the recording or sharing of content without consent^21^. For adolescents, problematic smartphone use interferes with academic responsibilities, social functioning, and psychological and physical wellbeing^22–24^.

Simultaneously, other research suggests no or only a small link between screentime and wellbeing in young people^25^.

In the UK, 90% of secondary schools now enforce restrictions or bans on smartphone use to reduce distractions and support classroom engagement^26^, although the efficacy of such measures is contested^27,28^.

To date, UK based evaluations of school smartphone policies have been limited. To extend emerging evidence, particularly by following pupils and schools prospectively through the implementation of a defined intervention, by combining individual level survey measures with school level disciplinary records collected from September 2023, and by examining heterogeneity across pupil subgroups, we aim to characterise the impact of a lockable smartphone pouch on educational attainment and behaviour, assess effects on general functioning and psychological wellbeing, and evaluate changes in school level indicators.

## METHODS AND ANALYSIS

### Overview

We will conduct a mixed-methods cohort study in UK secondary schools during the 2025-26 academic year, using retrospective and prospective data to evaluate the introduction of a lockable smartphone pouch.

### Ethics and Dissemination

This study was approved by the King’s College London Ethics Committee (Reference Number: HR-24/25-51368). To ensure data was process lawfully during this study, a Data Protection Impact Assessment (DPIA) was obtained on the 9^th^ of October 2025 and approved by the Northern Ireland Department of Education and King’s College London.

Participants under 16 will be confirmed to not have been opted out of the study by their parents prior to invitation to focus groups. All participants will give informed consent and will be instructed to keep the contents of their focus group or interview confidential. The researchers assured participants that if anything distressing will be mentioned, they could be signposted to support services.

A final report will be presented to the Northern Ireland Department of Education at study completion. The research will further generate multiple peer-reviewed publications covering the quantitative and qualitative components of the study, alongside presentations at relevant national and international conferences. Summary findings will additionally be shared with participating schools, pupils, parents, and policy makers in accessible formats.

### Patient and Public Involvement

Young people were involved in the design of the study questionnaire. A small group of adolescents in the target age range were consulted informally during questionnaire development to review item wording, clarity, and acceptability, and their feedback was used to refine the survey instrument prior to use in schools. Pupils were not formally involved in setting the research question, the overall study design, or recruitment.Findings will be shared with participating pupils and schools in accessible formats, and broader pupil and parent involvement in dissemination will be considered as the study progresses.

### Quantitative Methodology

#### Principle research objectives to be addressed

The primary objective is to assess the impact of the smartphone policy on educational attainment and behaviour.

The secondary objectives are to:

- Assess the impact of the smartphone policy on general functioning
- Assess the psychological impacts of the smartphone policy
- Assess the impact of the smartphone policy on subgroups who are most at risk
- Assess the impact of the smartphone policy on school-level measures

#### Outcomes

To assess the primary and secondary objectives, the below outcomes have been selected.

#### Primary outcomes

Educational attainment:

- Completion rate of homework

Behaviour:

- Total and domain scores on the Strength and Difficulties Questionnaire (SDQ)
- Frequency of classroom disruptions
- Participation in physical education
- Participation in extracurricular activities
- Breaktime interactions

#### Secondary outcomes

- Anxiety and low mood, measured using the Revised Child Anxiety and Depression Scale (RCADS) T-score
- Problematic smartphone usage, measured using the Smartphone Addiction Scale – Short Version (SAS-SV)
- Social media addiction, measured using the Bergen Social Media Addiction Scale (BSMAS)
- Smartphone usage measured by a series of questions relating to methods being implemented to reduce usage (i.e. parents taking phone, phone stacking, putting phone on silent) and their effectiveness

#### Aggregate School Data

The following outcomes will be collected from each school for every half term since September 2023:

- Exclusions
- Detentions
- CAMHS referrals
- School counsellor visits
- Parent visits due to disruption

### Study Design

The study follows a serial cross-sectional design. Students within schools participating in the introduction of a smartphone policy will be asked to complete a questionnaire up to three times: first to provide their thoughts prior to the smartphone policy implementation, and then in the following two terms after the initial questionnaire to provide their answers post the policy implementation.

Schools will also be asked to provide the aggregate level data for each half-term period since September 2023 until May 2026, alongside the date of smartphone policy implementation. The aggregate level data will be collected under an interrupted time-series design.

### Frequency and duration of follow-up

Primary and secondary measures will be collected from students at 0, 4 and 8 weeks (+/-2 weeks). Questionnaires will be kept open for at least 2 weeks at each timepoint to encourage as much data entry from students as possible.

### Measures

#### Demographics

- School
- School year
- Age
- Sex at birth
- Sex at birth same as gender
- Ethnicity
- Household yearly income
- Access to GP due to mental health/poor sleep
- Special educational needs (SEN)
- Autism Spectrum Disorder (ASD)
- Attention Deficit Hyperactivity Disorder (ADHD)
- Any other neurodivergent condition

#### Smartphone use

- Own a smartphone
- Current smartphone type
- Own more than one phone
- Own a simple mobile phone
- Age when given first phone

#### Primary outcomes

- Educational attainment measured by completion rate of homework (categorical)
- Behaviour measured using the SDQ – total score (scalar) and domains scores (scalar). Domains include emotional symptoms, conduct problems, hyperactivity, peer difficulties and prosocial
- Behaviour measured by frequency of classroom disruptions: how often they occur due to a smartphone (categorical), general frequency (categorical), involvement in a disruption due to a smartphone (categorical)
- Behaviour measured by frequency of PE participation (categorical) and engagement during PE lessons (categorical)
- Behaviour measured by involvement in extracurricular activities (binary) and frequency of extracurricular activities (categorical)
- Behaviour measured by frequency of interactions with friends during breaktime (categorical), quality of interactions (categorical), time spent during break talking to friends, parents, using WhatsApp, using other social media, on a smartphone in general and in detention (continuous)

#### Secondary outcomes

- Anxiety and depression measured using the RCADS – total, depression, and anxiety raw scores (scalar) and T-scores (scalar) adjusted for age and sex
- Problematic smartphone use measured using the SAS-SV – total score (scalar)
- Social media addiction measured using the BSMAS – total score (scalar)
- Smartphone usage measured by Reducing Usage Questions

The following data is to be collected at the school level, ideally by year group, for each half-term period from September 2023.

#### Aggregate school data

- Exclusions
- Detentions
- CAMHS referrals
- School counsellor visits
- Parent visits due to disruption

### Sample size estimation

To detect a reduction in symptoms of hyperactivity behaviour after the introduction of Odyssey pouch compared to the baseline score we will use a one-sample test. In order to detect a Cohen’s-d=0.2 (a small effect) with 90% power, type-1 error of 0.05 (two-sided test) and adjusting for clustering within school with an intra-cluster correlation (ICC) of 0.02 we would need a minimum of six schools and 400 participants per school (n=2,400). After accounting for a 25% loss to follow up (LTFU) of participants we need a minimum of 3,200 participants to enroll from at least six schools.

This is a conservative assumption. We would also have 90% to detect the same difference (d=0.2) with the same ICC (0.02), same LTFU and type-1 error, but if we only enrolled 134 participants we would achieve this in 8 schools (1,072 participants in total).

### Data Analysis Plan

#### Descriptives

A flow chart describing participant recruitment will be constructed. This will include the number of schools, number of participants consented at each timepoint and number of participants who completed the questionnaire at each timepoint.

Each of the outcome measures will be described at each timepoint. Means, standard deviations, or medians and interquartile ranges will be used for continuous variables. Frequencies and proportions will be used to describe categorical variables. Events will be described as the number occurred by each timepoint for aggregate school data. For outcomes such as the RCADS, SDQ, SAS-SV, results will be described using total scores and domain scores (continuous), as well as by clinically relevant cut-off values (categorical).

#### Inferential analyses

The analysis of student data will include all participants who have completed the outcome data, with the intent to estimate an intention-to-treat (ITT) type estimand. This will be estimated for the primary and secondary outcomes. All analyses will use a significance level=0.05.

#### Primary outcomes

All models will be adjusted for school, year group and any other variables (student-level or school-level) deemed relevant to the outcome. The primary outcomes will be analysed as follows:

#### Continuous outcomes

Continuous outcomes will be analysed with a linear regression model. Timepoint will be included as a categorical covariate. The adjusted mean difference for each timepoint will be presented with associated 95% confidence interval.

#### Binary outcomes

Binary outcomes will be analysed with a logistic regression model. Timepoint will be included as a categorical covariate. The adjusted odds ratio for each timepoint will be presented with associated 95% confidence interval.

#### Categorical outcomes

Outcomes with more than two categories will be collapsed into a binary framework and analysed as a binary outcome.

#### Secondary outcomes

Analysis of secondary outcomes will be consistent with the primary analysis.

#### Analysis of aggregate school data

The effect of the smartphone policy on the specified outcomes will be evaluated using an interrupted time series (ITS) design. The date of policy implementation defines the interruption point for the time-series analysis, and the outcome is a continuous variable representing the total number recorded in each school during each half-term period. The outcome will be modelled using a multilevel linear mixed model. Smartphone policy will be included as an indicator variable for before and after policy implementation. Time will be included as a continuous covariate to indicate each half-term period. A random intercept and slope over time will be fitted by school. Post-intervention time (i.e. continuous time from the start of the policy implementation) will be included. The model will be adjusted for school IMD, free school meal eligibility and other variables deemed relevant. If there are convergence issues, a simplified model including only a random intercept for school will be fitted

The analysis will estimate the baseline level and trend prior to policy implementation, the immediate level change in outcome following policy introduction, and the change in trend in the outcome after the policy (relative to the pre-policy trend). The adjusted mean differences will be presented alongside 95% confidence intervals.

### Sensitivity and subgroup analyses

The following subgroups will be investigated:

- SEN/neurodivergent diagnosis
- Index of multiple deprivation
- Smartphone implementation policy

Other subgroup and sensitivity analyses may be performed based on observations of descriptive data.

### Missing items in scales and subscales

The number (%) with complete data will be reported. If missing data guidance is provided for missing items within the scale scoring guide, this will be used. If no guidance is provided scales will be pro-rated for an individual if 20% or fewer items are missing. For example, in a scale with 10 items, prorating will be applied to individuals with 1 or 2 items missing. The average value for the 8 or 9 complete items will be calculated for that individual and used to replace the missing values. The scale score will be calculated based on the complete values and these replacements.

### Missing demographic data

If there is missing demographic data for any covariates, an approach such as including a missing indicator as a covariate in the model or simple mean imputation may be used.

### Method for handling multiple comparisons

No adjustment will be considered for multiple comparisons, allowing reviewers to make their own adjustment or estimates if they wish.

### Model assumption checks

The models assume normally distributed error terms (residuals); this will be checked following model fit and if these appear non-normal, transformations or bootstrapping of the standard errors (and subsequent confidence intervals) will be considered.

### Software

#### Data management

Qualtrics will be used to collect questionnaire responses from students. The platform is hosted by Qualtrics on secure cloud-based servers with access administered by King’s College London. Data will be provided in Excel format.

#### Statistical analysis

Stata (version 19.5 or higher) will be used for the main analyses. R (version 3.6.3 or higher) and R markdown may also be used for producing graphs and tables.

### Qualitative Methodology

#### Focus Groups

##### Participants

Students and teachers will be invited from each participating school to take part in focus groups during the autumn and spring terms of the 2025-2026 academic year. Student participants will either be selected by the school through student groups (e.g. student council) or by year group. Staff participants will self-select after being emailed an advertisement for the focus group from the school lead.

Eligibility criteria for students are: not opted out by their parents, if under the age of 16, and having a smartphone. Eligibility criteria for staff are that they are 18+ years of age and could speak English. No incentives will be provided to attendees of the focus groups.

##### Procedure and Materials

All participants will be provided with an information sheet prior to the focus group, and upon arrival complete a paper-based consent form. All consent forms will be collected by the researchers and later stored securely in a locked cabinet upon return from the schools. Participants will take part in an audio-recorded focus group with the researchers (JJ and AK) acting as facilitators. The focus groups will be conducted in person, in a private room in the schools, where participants will discuss their experiences of the new smartphone policy.

#### Semi-structured Interviews

##### Participants

School leads will be invited to semi-structured interviews to discuss their experiences of implementing the new smartphone policy.

##### Procedure

Interviews will take place online, via Microsoft Teams, where audio discussions will be recorded. School leads will be provided with an information sheet and will return an online e-consent form to the interviewer before starting the interview. The interviewer (NK) will be a separate member of the research team, who will not conduct the focus groups, and will have prior experience of conducting semi-structured interviews.

##### Data Processing

All transcripts will be processed and reviewed by JJ and AK. Each transcript will be checked against the focus group audio recording. All identifiable information will be redacted from the transcripts. All focus group participants will be given pseudonyms in the original transcripts that will later be replaced with anonymous participant numbers (e.g. P1, P2, P3). Upon completing transcript redactions and anonymisation, the transcript will be cross-checked by another researcher to confirm all identifiable information will be redacted and ready for analysis.

##### Data Analysis

Data will be analysed using Braun and Clarke’s^29^ six-phase approach to thematic analysis. This includes familiarisation with the data by reading the transcripts and fields notes, followed by initial coding using both inductive (i.e. derived from the data) and deductive approaches (i.e. informed by the topic guide and research aims). Codes will be organised into themes that reflect patterns in perceptions and experiences of the phone pouch intervention, with consideration given to similarities and differences between the stakeholder groups. Themes will be refined and finalised through consensus meetings with the expert advisory group and findings will be reported in relation to existing literature and implications for policy and practice

### Main guiding coding questions

#### Engagement with Learning

Does the use of mobile phone pouches affect attention, concentration, and engagement during lessons?

Are there impacts on academic performance (e.g., participation in discussions, assignment completion)?

Are there any other observed benefits or challenges related to student engagement? Participation in School Life:

- How does restricting phone usage affect students’ participation in extracurricular activities and school events?
- What changes occur in students’ social interactions during breaks and lunchtime?
- Are there other benefits or challenges identified?

#### Behaviour

- Does the use of mobile phone pouches reduce behaviour incidents linked to phone use?
- How does the intervention affect the classroom learning environment?
- Are there additional benefits or challenges in relation to student behaviour?

#### Wellbeing

- What impact does limiting phone access have on pupils’ mental and emotional wellbeing (e.g., anxiety, stress)?
- Do pupils report any significant changes in their wellbeing as a result of the pilot?
- What other benefits or challenges are related to pupils’ wellbeing?

Practicalities/ Implementation:

- What are the practical challenges of implementing mobile phone pouches (e.g., time spent on locking/unlocking pouches, enforcement issues)?
- Are there other practical considerations identified by schools?

## Data Availability

This is a protocol and no data is presently available

## Authors’ contributions

BC conceived the study and secured funding. BC and NK provided overall supervision of the project. ML developed the study database, with contributions from FM, JJ, and AK. FM developed the quantitative statistical analysis plan with input from BC. JJ and AK developed the qualitative analysis plan with input from CL and NK. ML, JJ, AK, and FM wrote the first draft of the manuscript. All authors contributed to revising the manuscript critically for important intellectual content and approved the final version for submission. BC is the guarantor.

